# Telomere shortening as a stress biomarker in children and adolescents affected by natural disasters

**DOI:** 10.1101/2021.02.20.21252124

**Authors:** Débora M. Miranda, Sabrina S. Magalhães, Daniela V. Rosa, Luiz A. De Marco, Jonas J. de Paula, Marco A. Romano-Silva

## Abstract

Natural disasters have a substantial psychosocial impact. A known biological marker of stress is telomere shortening. In this study, we tested the change in behavior symptoms and telomere length and its shortening in two-time points for about fifteen months, in populations that suffered extreme climate events, comprising flood or drought events. As expected, we observed telomere shortening in children and adolescents after a stressful situation which was directly associated with the worsening of externalizing symptoms and post-traumatic symptoms using a reliable change index. Beyond the psychosocial impact, natural events seem to affect the biology of individuals in development. These findings can help to understand vulnerabilities related to stress impact and to point target populations whose mitigation actions should be addressed in case of a disaster.

## 1. INTRODUCTION

Floods are the most common type of natural disaster worldwide [1]. Although floods are frequent disasters, droughts are long-term affecting large numbers of people. Drought is often unlimited in time since onset and end are sometimes hardly defined. It is also notably challenging for individuals to perceive and recognize it as a disaster, comprising a condition that affects chronic stress. Anyway, the incidence of these extreme weather events has increased by 46% since 2000 [2,3]. The interface of mental health and climate change is of significant concern because of increasing incidence and the fact that the psychological consequences of disasters exceed physical injury by 40:1 [3].

Mental health might be affected directly or indirectly. Direct effects involve the psychological trauma that followed the adverse experience, the destruction of landscapes which disturbs the sense of belonging and solace with the land [4]. The indirect effects occur by affecting physical health, aggravation of previous health problems, uncertain physical and social environment and infrastructure, caregiver burden that compromises the quality of care, compromising food and water shortages, influencing conflict and displacement, disturbing community wellbeing and loss of community identity, provoking school interruption, work disruption, and public service outages, apart from other consequences [3,4]. These indirect effects from a disaster might amplify the impact of a disaster.

The most common child behavioral symptoms related to direct affects on natural disasters are those related to post-traumatic stress [3,5]. After a disaster, children and adolescents present a higher risk of developing psychological distress when compared to adults [5]. Twenty-five percent of adolescents had faced a natural disaster in their lifetimes. For them, the vulnerability is associated with physiological and cognitive immaturity, limited physical skills, a higher metabolic rate that increases sensitivity to temperature, and dependency on others for care, protection, safety and provision [6,7]. These changes affect children and adolescents in the long term. The existence of critical windows of vulnerability during early childhood, when biological systems are developing, imposes another difficulty to approach these populations [6]. Stress might be a cumulative effect that impairs the ability to adapt and respond appropriately; and a burden to the systems at a critical developmental stage. Among the potential impairments related to exposure to chronic stress is the telomere shortening, depending on the individual cortisol exposure and reactivity [8–10]. In a recent meta-analysis, individuals recruited for psychological stress assessment were pooled and a small size effect was observed in telomere shortening, even after age adjustment [11]. Psychological stress and its cortisol responsiveness is crucial for children in the aftermath of a disaster, while the former may be more significant to chronic conditions [12]. Both are early adversity effects that impact biological features such as telomere shortening.

The intermediate and long-term periods after those events can last from months to years and are characterized by recovery post-event reconstruction [5]. To measure the effects of climate change related events, we propose to assess mental health reported behaviors and biological markers in two-time points, around 15 months apart from the first data collection. The chosen biological marker was a measure of telomere length and its attrition rate. At the tips of chromosomes, there are many TTAGGG sequence repeats. Telomere length shortens naturally during cell replication, but cellular and environmental stressors might accelerate it. The expected telomere length might be shortened and can follow behavior changes when an individual is affected by early stress events [13].

In this study, we observe behavioral symptoms and telomere shortening after an extreme weather event. The aim was to describe and correlate mental health problems with telomere shortening, approximately after a time-lapse of 15 months. Post-traumatic stress symptoms (PTSS) and general behavior were recorded and analyzed together with telomere length in the two-time points.

## 2. MATERIALS & METHODS

### Participants

We assessed 35 children and adolescents who were exposed to drought (n=19) or flood (n=16) (mean age=11.14+-3 years). The sample was a subsample of a previous study (Magalhães et al., 2020) on the consequences of extreme climate events on mental health.

All participants were assessed at two-time points. The assessment was forty days after the peak overflow, and the second one occurred after 14 months for flood conditions and after 17 months for drought. As we had observed in the previous study, post-traumatic symptoms of both conditions, here we pool them together to verify if the behavioral changes follow telomere shortening.

The participants affected by a flood event were from Rio Branco, a city in the eastern Amazon region in Brazil. It is a place with recurrent flood episodes, including a massive 2015 event, which was the most severe one the community faced, affecting over 21% of the population.

There are difficulties in identifying the beginning of a drought event. It was defined as a region under a period of below-average precipitation or above normal evaporation, having dryness as a result. We recruited children and adolescents from the town of Francisco Sá, in the semi-arid zone of the state of Minas Gerais in Brazil, which is an area affected by drought [14].

Besides living in the respective chosen locations, other inclusion criteria were a minimum age of 6 years, written consent by a parent or caregiver, written assent by the child, and completion of the pre- and post-assessment of all the instruments below. We excluded subjects with a previous diagnosis of epilepsy, seizures, or other neurological diseases. The study followed the Declaration of Helsinki, and the local ethics committee approved all procedures of this study under the registration number CAAE: 26886814.9.0000.5149.

### Psychological assessment and data modeling

We assessed internalizing and externalizing symptoms using the Brazilian version of the Child-Behavior Checklist for 6-18 years (CBCL6-18) [15]. This measure comprises 113 questions about different aspects of child-behavior, presenting a dimensional approach to psychological symptoms. Parents and caregivers filled the scale. We adopted the Externalizing and Internalizing symptoms scores in the present study. The age and cultural background influence CBCL 6-18 scores, so we used scaled T-scores controlling for these factors. Also, the Brazilian version of the Children’s Revised Impact of Event Scale (CRIES-8) was used to document post-traumatic stress consequences of the exposure to drought or flood [16]. To test the potential age effects on CRIES-8 reporting, the association between age and CRIES was evaluated. However, we found no correlation between the total score and the participant’s age (r= -0.148, p=0.363), then we adopted in subsequent analysis its raw-scores.

A brief comparison of pre and post-assessments showed a very heterogeneous pattern of change, with participants showing improvement, worsening, or stabilization of CBCL 6-18 and CRIES-8 scores (Figure 1). Descriptively, as a group, we observed minor changes for CBCL 6-18 scores and more substantial changes in CRIES-8, despite high individual variability. The scores in CBCL 6-18 and CRIES-8 may also change between two-time points because of measurement error, although the test usually shows high reliability (internal consistency >0.75).

**Figure 1:**
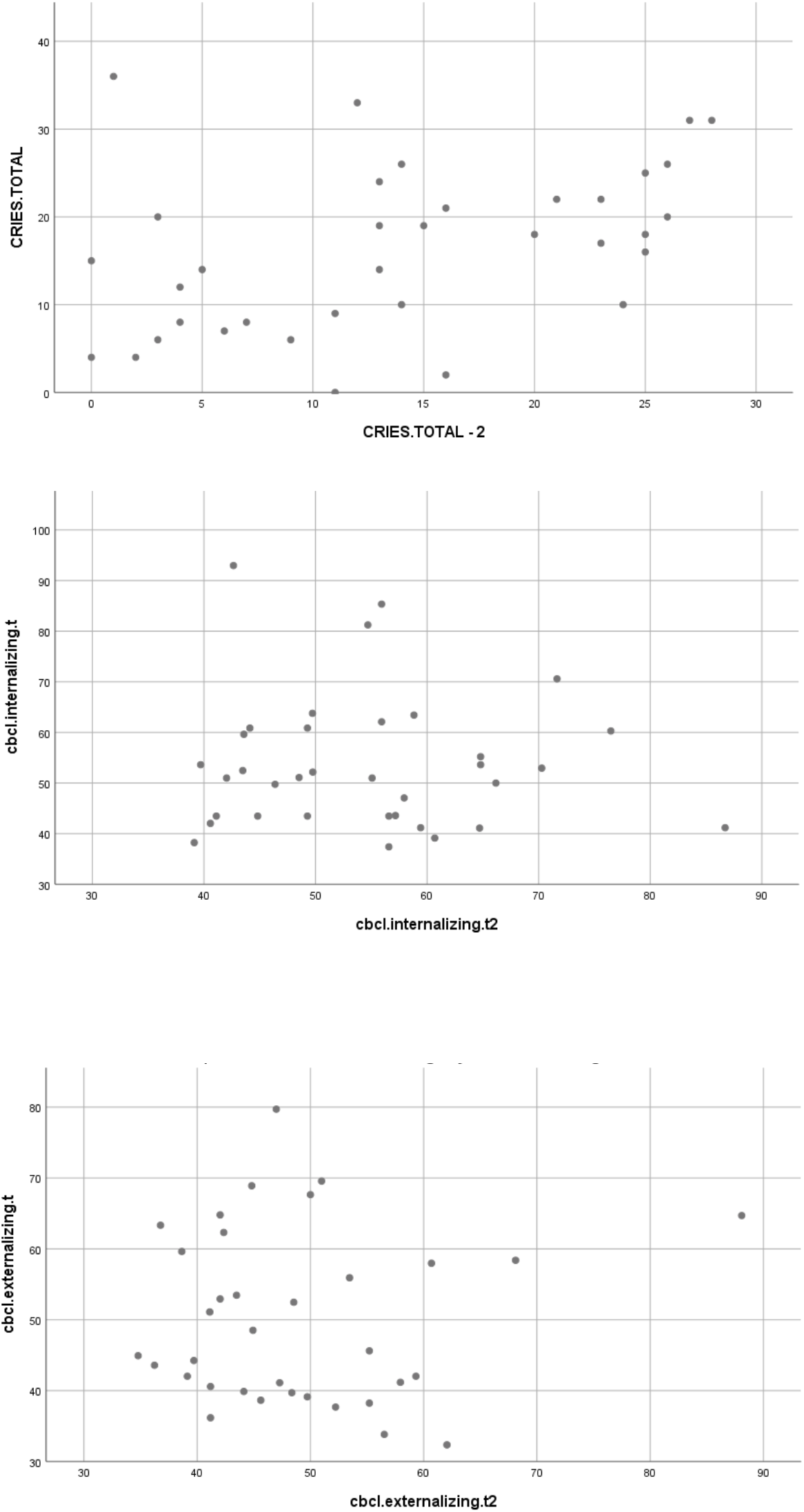
individual CBCL 6-18 and CRIES-8 scores in baseline (vertical axis) and follow-up (horizontal axis). CBCL: Child-Behavior Checklist, CRIES: Children’s Revised Impact of Event Scale.

Since we found a very heterogeneous pattern of change in psychological measures and to avoid the latter bias of test-retest reliability, we adopted an individual case approach, computing reliable change statistics. This procedure allows individual comparisons of pre-post-test scores to be corrected for test reliability and measurement errors. Reliable Change Indices (RCI) were computed for each subject in CBCL 6-18 and CRIES-8 measures and transformed into Z-Scores. We adjusted these results so that higher RCI values represented a reliable worsening of psychological symptoms.

### Telomere length assay

Peripheral blood samples were collected in tubes containing EDTA, followed by DNA extraction with a high salt method (Lahiri and Schnabel, 1993). The DNA was quantified using a NanoDrop Spectrophotometer Thermo Scientific, Nanodrop 200 model, and diluted to 75ng in 96 well plates. The relative quantification method, described by Cawthon (2002), was used to measure telomere length. The telomere reaction proceeds for one cycle at 95°C for 10 min, followed by 18 cycles at 95°C for 15 s and 54°C for 2 min and primers used were Tel-1 primer (GGT TTT TGA GGG TGA GGG TGA GGG TGA GGG TGA GGG T) and Tel-2 primer (TCC CGA CTA TCC CTA TCC CTA TCC CTA TCC CTA TCC CTA). The 36B4 reaction proceeded for one cycle at 95°C for 10 min, followed by 30 cycles at 95°C for 15s and 58°C for 1 min 10 s and primers used were 36B4u (CAG CAA GTG GGA AGG TGT AAT CC), 36B4d (CCC ATT CTA TCA TCA ACG GGT ACA A). For PCR reactions, PlatinumTaq (Invitrogen) was used, and amplicon formation was monitored using SYBR-Green fluorescent dye (Invitrogen). All PCR reactions and fluorescence measures were carried out in an ABI-7500 real-time PCR machine (ABI). The reaction was performed in triplicate for each sample, and results were averaged in further calculations. For telomere length quantification, cycle thresholds (Ct) for each telomere and control gene 36B4 PCR reaction were calculated using the ABI software algorithm. The telomere/control gene 36B4 (T/C) ratio reflects the relative size of telomere for each sample. Considering the exponential kinetics of the PCR reaction, this ratio may be expressed as the following equation: 2-ΔCt, where –ΔCt=-(Ct telomere - Ct control gene 36B4) of sample n. For group comparisons, 2-ΔCt values for each sample were grouped and analyzed together.

### Statistical analysis

The telomere attrition index was calculated using the ratio between the last telomere measure and the first one. It was log-transformed (base 2) to stress the differences found and to facilitate interpretation since positive values show telomere growth and negative ones, meaning telomere reduction between the two assessments. The comparison between baseline and follow-up telomere length was conducted using paired samples, t-tests and effect sizes computed using Cohen’s d for within-subjects design. We tested the association between RCI measures for CBCL 6-18 and CRIES-8 scores and the type of adverse event (flood or drought) using general linear models in SPSS 22.0 (IBM Inc, 2014).

## 3. RESULTS

We found a reduction in telomere length between the first and second evaluation (t=4.86, p<0.001) with a moderate effect size (d=0.360), which was expected since telomere length usually decreases. Table 2 shows the General Linear Model results. The overall model was significant (F=4.50, p=0.006, pn2=0.37), and the reliable increase of Externalizing Symptoms (F=5.80, p=0.022, pn2=0.16) was associated with higher telomere shortening, while a constant improvement in CRIES-8 scores was associated with lower telomere shortening (F=6.15, p=0.019, pn2=0.17). Reliable change in internalizing symptoms (p=0.575) and the type of stressful event was not associated with telomere shortening (p=0.920). Trajectory of telomere length values from time point 1 to time point 2 were shown in Figure 2.

**Table 1:** Participants data

|  | 40 days after event |  | Follow-Up |  |
| --- | --- | --- | --- | --- |
|  | M | SD | M | SD |
| Age | 11.14 | 3.00 | 12.40 | 3.51 |
| Telomere Length | 220.63 | 155.18 | 173.57 | 133.17 |
| CBCL 6-18 Internalizing Symptoms | 53.68 | 13.26 | 54.53 | 11.37 |
| CBCL 6-18 Externalizing Symptoms | 50.07 | 12.12 | 48.82 | 10.56 |
| CRIES-8 | 16.37 | 9.28 | 13.86 | 9.01 |

CBCL: Child-Behavior Checklist, CRIES: Children's Revised Impact of Event Scale, M=Mean, SD: Standard-Deviation

**Table 2:** Factors associated with higher telomere attrition (analyzed by general linear model)

| Predictor | F | P | $\eta_p^2$ |
| --- | --- | --- | --- |
| Reliable increase of CBCL 6-18 Internalizing symptoms | 0.32 | 0.575 | - |
| Reliable increase of CBCL 6-18 externalizing symptoms | 5.80 | 0.022 | 0.16 |
| Reliable decrease of CRIES-8 symptoms | 6.15 | 0.019 | 0.17 |
| Adverse condition (flood x drought) | 0.01 | 0.920 | - |

CBCL: Child-Behavior Checklist, CRIES: Children's Revised Impact of Event Scale, $\eta_p^2$ = partial *eta* squared.

## 4. DISCUSSION

Drought causes changes in the economic structure, following alterations in food and water provisions, and land degradation. Long-term drought has been associated with conflict and forced migration [17], which were also linked to adverse psychosocial outcomes [18]. The domestic routine is disrupted and becomes a daily challenge. In addition, adversities and mental health symptoms are accumulated, such as psychological distress, depression, and generalized anxiety [19]. Flood events caused disturbances for a short time and resulted in the recovery and restoration of community functioning [5]. Our follow-up occurred during the fifteen months post-event when telomere shortening was observed in both conditions. Apart from the differences in the effects of the two stressful events, in this phase and under chronic stress, the telomere attrition rate behaved in the same way.

Children and adolescents answer to stress with behavior changes and telomere shortening. Most neuropsychiatric disorders emerge from complex interactions between stressors, vulnerabilities and compensatory mechanisms, resulting in abnormal developmental trajectories. Youth exposed to disasters present a higher risk for developing an extensive set of responses, including PTSS, depression, anxiety, phobias, sleep disorders, attachment disorders, aggression, functional impairment, substance use, suicidal ideation and behavior, risk-taking behaviors, and physical health problems [20–23]. A meta-analysis conducted by Alisic et al. (2014)[24] and our previous data showed PTSS, depression, and anxiety in the aftermath of the event.

Stress may change the response of the organism, hence establishing a vulnerability condition where mental health problems can emerge. Telomere length variation seems to happen mostly in the first years of life [25] being moderated by environmental characteristics. In this study, we expected telomere shortening, consequent cellular aging, and behavior changes caused by early exposure to adversity [13]. In a previous meta-analysis, there was an observation of the association between events causing post-traumatic disorders in children, resulting in telomere shortening [26]. In our data, we observed a tendency to have a shortening in telomere length. A greater impact on telomere shortening was observed in those who had more symptoms and important reduction in PTSS scores. Therefore, subjects with higher behavioral recovery showed a higher biological impact of stress. It might reflect that they were more affected at first, resulting in more symptoms just after the disaster. However, mitigation actions and resilience resulted in behavioral improvement with a biological cost dependent on environmental sensitivity. This vulnerability represents the result of interaction between risk and protective factors with the individual susceptibility, and it determines whether an individual exhibits adverse mental health outcomes in response to an extreme event [27]. The possibility of a third telomere prospective evaluation would show if the organism recovers after the initial stress, but it is maintained under allostatic load and sped up telomere loss.

Internalizing and externalizing problems have been frequently associated with changes in telomere shortening in childhood [28,29]. Here, we tested the dynamic of internalizing and externalizing symptoms and telomere shortening after a stressful climate-related event. The relationship between psychological symptoms and telomere shortening is expected. Shorter telomeres were observed in mothers with postpartum depression, and it was as a predictive feature of the presence of internalizing and externalizing symptoms in children, suggesting the stressful features lived by mothers are underlying the behavioral problems in children [30]. The findings on externalizing and internalizing symptoms deserve a careful evaluation of the stress response and subsequent biological changes, in both conditions, to have a clear picture concerning their impacts.

The differential susceptibility model might help to understand why children with increasing symptoms of externalizing disorders seem to have greater telomere attrition [31]. Externalizing behaviors are common forms of childhood maladjustment, and it was recognized as a major risk factor for psychopathology in adulthood. In a study with adolescents, common genetic features and liability contributed to the co-occurrence of depression, conduct, and hyperactivity symptoms [29]. Based on twin studies, there are common grounds to say that comorbidities have genetic features [29,32]. Children with externalizing symptoms are more vulnerable to harmful effects related to stressful events. Under stressful conditions they suffer more impact shown as telomere shortening. The shortening of telomere reflects a stressful condition large enough to cause biological stress. However, the underlying mechanisms for this biological impact are still not understood. Thus, those kids should be the target of public policies, since they are definitely the most reactive and impacted by stressful conditions, and the ones under highest biological impact. Their resilience is less rooted, showing a clear need for a more supportive environment to buffer the impacts of stress promoted by extreme climate events.

There are some limitations to our study due to the biological complexity and social information. Stress hormones induced immune cell mobilization changes in certain hormonal and maturational conditions [33]. Depending on the phase of stress response and the type of cells mobilized, the telomere length might differ [33,34]. We compared the responses at the same time after the event, but it was not so clear the beginning of the drought condition. After 15 months, for both conditions, there was a chronic stress effect. In further studies, hormonal and cell type characterization might help to clarify the differences related to long-term stress and the effect on telomere shortening. We did not test the mitigation actions performed at each spot, so we cannot infer about it. We re-genotyped the whole sample to ensure that the telomeric size was reliable. It is important to emphasize that telomere length error is related to sampling bias, and in this study, we expected population segregation, since both regions are known for the constant migration. However, as the variability related to the event should be the same, we calculated the attrition rate instead of the length to have more accurate data.

In conclusion, our results add evidence to the considerable impact of extreme climate events on children and adolescents, with potential long-term effects on their physical and mental health. This must be considered in mitigation plans when approaching the affected populations.

## Data Availability

The data that support the findings of this study are available on request from the corresponding author,MAR-S. The data are not publicly available due to restrictions, their containing information that could compromise the privacy of research participants.

## Acknowledgements

This study was funded by grants from the governmental agencies CAPES, CNPq and FAPEMIG. DMM, LADM and MAR-S are CNPq research fellows.

